# Effectiveness and cost-effectiveness of orthopaedic modifications to off-the-shelf footwear for people with first metatarsophalangeal joint osteoarthritis: study protocol for a randomised controlled trial

**DOI:** 10.64898/2026.05.12.26352874

**Authors:** Sjanne Veenstra, Chantal M. Hulshof, Judith E. Bosmans, Dieuwke Schiphof, Margot van der Grinten, Sabine E. Kloprogge, Carla Braam, Leen Nugteren, Sita M.A. Bierma-Zeinstra, Marienke van Middelkoop

## Abstract

**Introduction:** Osteoarthritis (OA) is a chronic joint disease, often leading to pain, joint stiffness and impaired function. The first metatarsophalangeal (MTP-1) joint is the most frequently affected joint in foot OA. Footwear interventions might have potential to reduce pain for people with MTP-1 joint OA. The aim of this study is to determine the effectiveness and cost-effectiveness of orthopaedic modifications to off-the-shelf footwear in addition to usual care, compared to usual care alone, for people with MTP-1 joint OA.

**Methods and analyses:** We perform a pragmatic, non-blinded, two-armed, parallel-group, randomised controlled trial (RCT). A total of 136 people with MTP-1 joint OA and presence of foot pain are recruited. Participants are randomised to orthopaedic modifications to off-the-shelf footwear in addition to usual care or to usual care alone. The footwear modifications comprise a combination of sole-stiffening, rocker sole adjustments and custom-made insoles. During a 12-month follow-up period, participants receive monthly questionnaires. Primary outcomes include walking pain at 6-month follow-up and quality-adjusted life years and societal costs at 12-month follow-up. Secondary outcomes include walking pain at 12-month follow-up and foot health, physical activity level, patient acceptability and self-reported recovery at 6- and 12-month follow-up. Intention-to-treat and per-protocol analyses will be performed using (generalised) linear mixed models.

**Ethics and dissemination:** The study is approved by the local Medical Ethics Committee of the Erasmus MC University Medical Center Rotterdam, The Netherlands (MEC-2024-0615). Prior to study participation, participants provide informed consent. Results will be disseminated amongst researchers through peer-reviewed scientific articles and presentations at conferences; and amongst people with MTP-1 joint OA and healthcare professionals through layman articles in newsletters, on websites and on social media.

**Discussion:** This is the first RCT to investigate the effectiveness and cost-effectiveness of orthopaedic modifications to off-the-shelf footwear in addition to usual care, compared to usual care alone for people with MTP-1 joint OA. Study findings will support healthcare professionals in making substantiated decisions in the treatment of people with MTP-1 joint OA.

**Trial registration number:** Overview of Medical Research in the Netherlands (OMON): NL87646.078.24

## INTRODUCTION

Osteoarthritis (OA) is a chronic whole-joint disease that causes pain, joint stiffness and impaired function, leading to a high individual burden and reduced quality of life (1, 2). Additionally, OA imposes a substantial societal burden on economies, through direct costs for disease management, such as hospitalisation, outpatient visits and medication, as well as indirect costs arising from productivity losses (3). The global prevalence of OA increased from 256 million in 1990 to 595 million in 2020 and is expected to continue to increase due to the ageing global population and the rising prevalence of obesity (1, 4). To limit the individual and societal burden of OA, research into effective and cost-effective treatments for OA is highly necessary. Until now, research to OA has predominantly focused on the joints of the knee, hips and hands, while foot OA has been highly underresearched (5).

The population prevalence of symptomatic radiographic foot OA is estimated at 17% in adults aged 50 years or older and increases with age (6). The first metatarsophalangeal (MTP-1) joint is the most frequently affected joint in foot OA, with an estimated population prevalence of 8% in adults aged 50 years or older (6). Several clinical characteristics are associated with radiographic MTP-1 joint OA severity, including dorsal hallux and MTP-1 joint pain, hallux valgus, keratotic lesions on the dorsal side of the hallux and MTP-1 joint; and reduced MTP-1 joint dorsiflexion range of motion (7). Thereby, people with symptomatic and radiographic MTP-1 joint OA experience more foot pain, reduced physical functioning and reduced foot-specific health-related quality of life, compared to people without MTP-1 joint OA (8). Effective treatment strategies are needed to improve patient outcomes and reduce the burden on people with MTP-1 joint OA.

For the treatment of OA in general, non-surgical and non-pharmacological treatment strategies in early stages of the disease are recommended, requiring proactive management of general practitioners from the first consultation (9). Whereas international guidelines exist for the treatment of OA in the hand, knee and hip (10–12), guidelines for the treatment of MTP-1 joint OA are lacking. Management of MTP-1 joint OA in general practice in The Netherlands predominantly includes explanation, advice on footwear, referrals to an orthopaedic surgeon or podiatrist, advice to use paracetamol and prescription of non-steroidal anti-inflammatory drugs (NSAIDs) (13). Management of MTP-1 joint OA by podiatrists in Australia and the United Kingdom predominantly includes taping or padding and footwear modifications as well as advice on pacing activities, weight loss, footwear, and the use of heat, and the advice to discuss the use of pain medication with a general practitioner (14). Despite the wide range of available treatment strategies to approach MTP-1 joint OA in primary care, evidence on the effectiveness and cost-effectiveness of non-surgical treatments for MTP-1 joint OA is limited (15).

Biomechanical studies showed that footwear modifications can reduce MTP-1 joint peak pressure (16) and MTP-1 joint maximum dorsiflexion (17, 18) during walking, suggesting that footwear modifications might have potential to reduce walking pain for people with MTP-1 joint OA. One clinical trial showed that both foot orthoses and rocker sole footwear induce similar and clinically relevant foot pain reductions in people with MTP-1 joint OA (19). However, other clinical trials showed that both stiffening insoles and foot orthoses do not result in clinically relevant foot pain reductions in people with MTP-1 joint OA, compared to sham insoles (18, 20). Until now, only one feasibility study compared a footwear intervention to usual care in the treatment of people with MTP-1 joint OA (21). Whereas the study showed that it is feasible to conduct a clinical trial, comparing a podiatry intervention, including foot orthoses, exercise, manual therapy, and shoe advice, to usual care delivered by a general practitioner, a larger, powered follow-up clinical trial has not yet been conducted. Furthermore, the effectiveness and cost-effectiveness of a combination of multiple footwear modifications (i.e. a combination of sole stiffening, rocker sole adjustments and custom-made insoles) in the treatment of MTP-1 joint OA have never been studied. This research gap has also been acknowledged by the International Foot and Ankle Osteoarthritis Consortium, which highlighted the need for further research on footwear and orthoses in the treatment of foot and ankle OA (22).

Therefore, we perform a randomised controlled trial with the aim to determine the effectiveness and cost-effectiveness of a combination of multiple orthopaedic modifications to off-the-shelf footwear in addition to usual care, compared to usual care alone, for people with MTP-1 joint OA in primary care. Additionally, we perform a qualitative interview study with the aim to identify barriers and facilitators experienced by people with MTP-1 joint OA and healthcare professionals regarding orthopaedic modifications to off-the-shelf footwear as treatment of MTP-1 joint OA in primary care.

## METHODOLOGY

### Study design

This is a pragmatic, non-blinded, two-armed, parallel-group RCT. Participant enrollment began in June 2025 and ended in February 2026. Participants are randomised to either (1) orthopaedic modifications to off-the-shelf footwear in addition to usual care or (2) usual care alone (Figure 1). Considering the 12-month follow-up, data collection will take until February 2027. This study protocol follows the 2025 SPIRIT updated guidelines for protocols on randomised trials (23).

**Figure 1:**
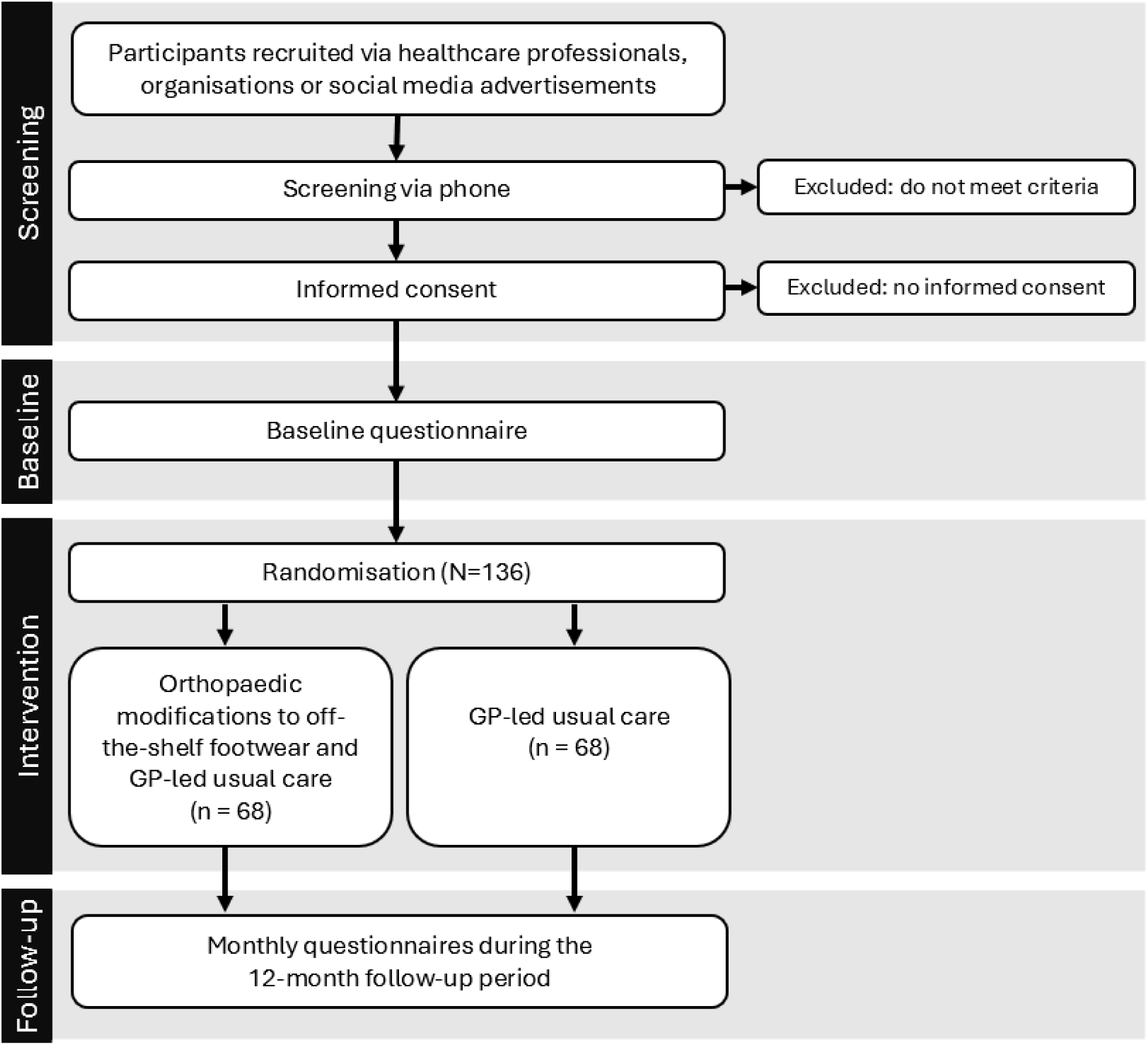
Flow diagram of study phases.

### Participants

People with MTP-1 joint OA are recruited through healthcare professionals, organisations and advertisements. Healthcare professionals such as general practitioners, podiatrists, medical pedicures and physiotherapists are asked to invite people with MTP-1 joint complaints to sign up for this study. In addition, organisations and associations concerning OA, general health or footwear are asked to inform their members or audience about the study through social media posts or articles on their websites or in newsletters. Finally, newspaper articles and advertisements on social media are used to recruit participants in the general population. People who signed up for the study are screened by telephone on the inclusion and exclusion criteria (Table 1). If eligible for study participation, they receive an information package and consent form via post or e-mail.

**Table 1:**
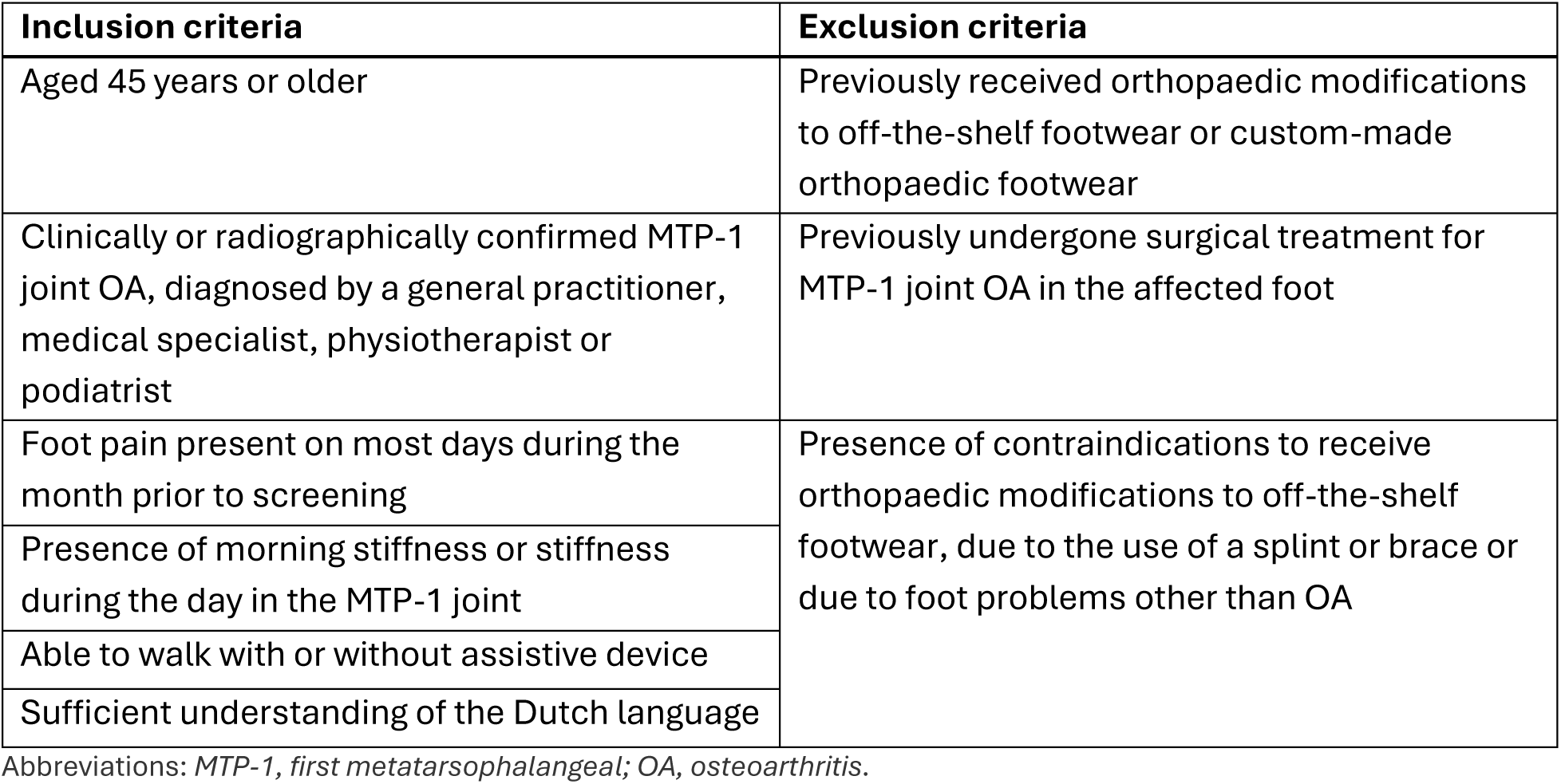
Inclusion and exclusion criteria for study participation.

| Inclusion criteria | Exclusion criteria |
| --- | --- |
| Aged 45 years or older | Previously received orthopaedic modifications to off-the-shelf footwear or custom-made orthopaedic footwear |
| Clinically or radiographically confirmed MTP-1 joint OA, diagnosed by a general practitioner, medical specialist, physiotherapist or podiatrist | Previously undergone surgical treatment for MTP-1 joint OA in the affected foot |
| Foot pain present on most days during the month prior to screening | Presence of contraindications to receive orthopaedic modifications to off-the-shelf footwear, due to the use of a splint or brace or due to foot problems other than OA |
| Presence of morning stiffness or stiffness during the day in the MTP-1 joint |  |
| Able to walk with or without assistive device |  |
| Sufficient understanding of the Dutch language |  |

Abbreviations: MTP-1, first metatarsophalangeal; OA, osteoarthritis.

Participants who meet all criteria and are willing to participate, are asked to provide informed consent, either written or digitally. After providing consent, participants receive the baseline questionnaire, followed by monthly follow-up questionnaires (see paragraph ‘Data collection’).

### Randomisation

After completion of the baseline questionnaire, participants are randomised into the intervention or control group with an allocation ratio of 1:1. Block randomisation (random size 4 or 6), stratified by symptom duration (> 2 years versus ≤ 2 years) is employed using the electronic data capture (EDC) system Castor (Castor EDC, Ciwit B.V., Amsterdam, the Netherlands). Blinding of participants, researchers and healthcare professionals is not possible, considering the nature of the intervention. Researchers inform participants on their allocation outcome via a telephone call. Additionally, general practitioners of participants are informed on their study participation and allocation via telephone (intervention group) or post (control group). Participants allocated to the control group are allowed to cross-over to the intervention group after 6 months of study participation on their own initiative, because a previous interview study from our group showed that this would increase willingness amongst people with MTP-1 joint OA to participate in the trial (24).

### Intervention

#### Orthopaedic modifications to off-the-shelf footwear

Participants allocated to the intervention group receive orthopaedic modifications to one pair of their own off-the-shelf footwear, provided by orthopaedic shoe technicians (pedorthists).

When the researchers inform participants in the intervention group on their allocation outcome, they will additionally ask them to hand in footwear that they regularly wear and to wear the modified footwear in their daily life. Participants will receive an information sheet via e-mail, describing the criteria that footwear should meet in order to be suitable for orthopaedic modifications (i.e. sufficient length and width, adjustable instep, removable insole, firm heel counter, and firm midsole). They are instructed to contact the researchers or consult the orthopaedic shoe technician if they are unsure if their shoes are suitable for orthopaedic modifications.

Participants visit an orthopaedic shoe technician two or three times at a location of their preference in the Netherlands. During the first consultation, participants hand in their off-the-shelf footwear and the orthopaedic shoe technician assesses foot posture, MTP-1 joint range of motion and pain symptoms to determine which orthopaedic modifications are required. Subsequently, the off-the-shelf footwear along with the instructions on the required modifications are sent to a single central production site in the Netherlands, where shoemakers who are specialised in applying orthopaedic modifications to off-the-shelf footwear, provide the required modifications to the footwear. The modified footwear gets returned to the visiting location. During the second consultation, usually three weeks after the first consultation, the orthopaedic shoe technician hands over the modified footwear to the participant. The orthopaedic shoe technician additionally provides instructions on the optimal duration to wear the modified footwear, starting with one hour per day and gradually building up to wearing them all day after five consecutive days. This allows participants to get used to the modifications in order to prevent pain and discomfort in the subsequent weeks. During the third consultation, generally six weeks after the second consultation, participants are contacted by the orthopaedic shoe technician by phone to verify whether they are satisfied with the modified footwear. If participants are not satisfied with their modified footwear, they can visit the orthopaedic shoe technician again to discuss possible adjustments.

The orthopaedic modifications can include a combination of sole stiffening and rocker sole adjustments; custom-made insoles and additional modifications (Figure 2A). For sole stiffening, the outer sole is detached from the midsole and a carbon foot plate (thickness 2.5 mm, hardness shore 50) is glued underneath the midsole, aiming to reduce dorsiflexion of the MTP-1 joint (17, 18). For rocker sole adjustments an ethylene-vinyl acetate (EVA) midsole (hardness shore 50, variable thicknesses) is glued underneath the carbon foot plate that was glued underneath the original midsole of the footwear, and sanded into the shape that is needed to obtain the desired rocker sole parameters. These parameters include sole thickness and toe spring, as well as forefoot and heel apex position and forefoot apex angle (Figure 2B). The aim of rocker sole adjustments is to optimize foot motion and redistribute foot loading during the stance phase (i.e. from heel strike to toe-off) and stimulate early offloading of the forefoot, thereby reducing MTP-1 joint motion and subsequently reducing the peak pressure on the MTP-1 joint during walking (16). After applying and sanding the EVA midsole, the original outer sole is reattached underneath.

**Figure 2:**
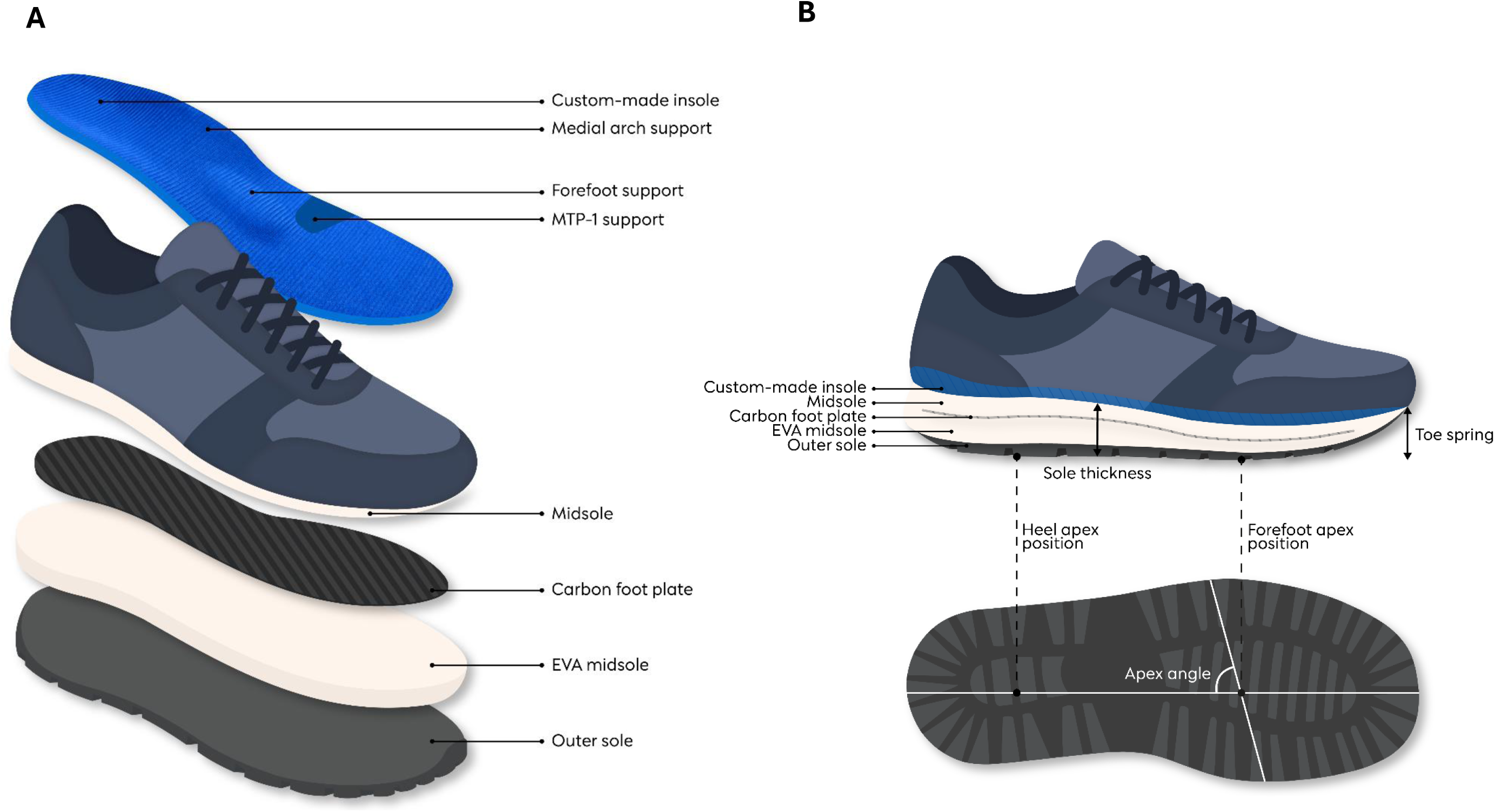
The main orthopaedic modifications that are applied to off-the-shelf footwear of people with MTP-1 joint OA, including a custom-made insole to provide medial arch support, forefoot support and MTP-1 support; a carbon foot plate to reduce MTP-1 joint dorsiflexion; and an EVA midsole to optimize rocker sole parameters (A). Rocker sole parameters include sole thickness, toe spring, forefoot and heel apex position and forefoot apex angle (B). Abbreviations: *EVA, ethylene-vinyl acetate; MTP-1, first metatarsophalangeal*.

Foot orthoses consist of a combination of elements (thicknesses of 3, 6, or 9 mm), depending on the person’s needs. Elements that are specific for MTP-1 joint OA include medial arch support, forefoot support (i.e. drop-shaped pelotte, T-shaped pelotte or transverse arch support) and MTP-1 support (i.e. elevation of the MTP-1 area of the orthoses), with the aim to create space within the MTP-1 joint and limit MTP-1 joint compression (16). Additional modifications to the shaft or heel of the footwear include replacement of the closure, adding textile laminates between the upper and lining material of the shaft or tongue, shifting the foot opening forward, stretching or widening the shaft and lowering or elevating the heel. The orthopaedic modifications result in minimal changes to the external appearance of the footwear.

#### Usual care

All participants receive usual care, which may include explanation, advice on footwear, referrals, advice to use paracetamol and prescription of NSAIDs (13). Since guidelines for the treatment of MTP-1 joint OA are lacking, usual care will vary amongst participants. The usual care that participants receive during follow-up, is on their own or healthcare providers’ initiative and is documented by self-report in questionnaires.

#### Co-interventions

Participants allocated to the control group cannot be referred to an orthopaedic shoe technician for delivery of orthopaedic modifications to off-the-shelf footwear or custom-made orthopaedic footwear in the first 6 months of study participation.

### Data collection

Participants are assessed at 13 time points (T) in the 12-month study period. After the baseline questionnaire (T0), participants receive monthly follow-up questionnaires (T1-T12). These will take either 60 minutes (T0, T3, T6, T9, T12) or 5 minutes (T1, T2, T4, T5, T7, T8, T10, T11) to complete. People with bilateral MTP-1 joint OA are asked to answer questions on foot pain regarding their most painful foot. The questionnaires are sent digitally by Castor EDC and reminders are sent automatically after 3 and 6 days. If participants have not completed the questionnaire after 10 days, the researchers contact them via e-mail or phone and ask them to complete the questionnaire.

### Outcome measures

#### Primary outcome measures

Primary outcome measures include walking pain on an 11-point Numeric Rating Scale (NRS) at 6-month follow-up and Quality-Adjusted Life-Years (QALYs), measured with the five-level version of the EQ-5D questionnaire (EQ-5D-5L) and societal costs, measured with the iMTA Medical Consumption Questionnaire (iMCQ) and iMTA Productivity Cost Questionnaire (iPCQ) at 12-month follow-up (Table 2).

**Table 2:** Overview of descriptive variables and outcome measures collected by questionnaires during the 12-month study period.

| Domain | Measurement variable | Timepoint |
| --- | --- | --- |
| **Descriptive variables** |  |  |
| *Demographics* | Body weight and height | 0, 3, 6, 9, 12 |
|  | Age, sex, menopausal status, education level and work status | 0 |
| *Lifestyle* | Current use of alcohol and tobacco | 0 |
|  | Sport participation in the past 10 years (type of sport, duration and frequency) | 0 |
|  | OSPAQ: occupational time spent sitting, standing, walking and doing heavy labour (25) | 0 |
| *MTP-1 joint OA-specific descriptive data* | Site of MTP-1 joint OA and symptom duration | 0 |
|  | Morning stiffness in MTP-1 joint | 0, 3, 6, 9, 12 |
|  | Previous treatments and medication use and their perceived effect on complaints on a 7-point Likert scale | 0 |
|  | Type of footwear predominantly worn | All |
|  | Current use of insoles and assistive devices for walking | 0 |
| *General health status* | SF-12: physical and mental health (26) | 0 |
|  | SCQ: comorbidities (27) | 0 |
|  | Presence of OA in other joints and presence of foot diseases other than OA | 0 |
| *Trust in the intervention* | Expectation on the ability of orthopaedic footwear modifications to off-the-shelf footwear to reduce walking pain | 0 |
| *Characteristics needed for behavioral change* | 6-item COM questionnaire: physical and psychological capability; physical and social opportunity; and reflective and automatic motivation to wear modified footwear (28) | 0 |

| **Primary outcomes** |  |  |
| --- | --- | --- |
| *Walking pain* | Foot pain during walking on an 11-point NRS | All |
| *Quality-adjusted Life Years* | EQ-5D-5L: health status on mobility, self-care, daily activities, pain/discomfort and anxiety/depression (29) | 0, 3, 6, 9, 12 |
| *Societal costs* | iMCQ: healthcare use and patient and family costs related to MTP-1 joint OA (30) | 3, 6, 9, 12 |
|  | iPCQ: productivity loss due to absence from paid work related to MTP-1 joint OA (31) | 3, 6, 9, 12 |
| **Secondary outcomes** |  |  |
| *Foot pain at rest and during physical activity* | Foot pain at rest, during physical activity in general and during the type of physical activity that is reported as most painful on an 11-point NRS | All |
| *Self-reported recovery<sup>§</sup>* | GROC: overall change in complaints over the past 3 months on a 7-point Likert scale (32) | 3, 6, 9, 12 |
| *Foot health* | FHSQ: foot pain, foot function, footwear and general foot health (33) | 0, 3, 6, 9, 12 |
| *Physical activity level* | SQUASH: physical activity in commuting, work, leisure time and household activities (34) | 0, 6, 12 |
|  | IPAQ-SF: time spent on vigorous- and moderate-intensity physical activity, walking and sitting (35) | 0, 3, 6, 9, 12 |
| *Patient acceptability* | PASS: patient's acceptability of current health status (36) | All |
| **Other outcomes** |  |  |
| *Current treatments* | Current treatments for MTP-1 joint OA | 0, 3, 6, 9, 12 |
| *Adherence to the intervention\** | Frequency and duration of wearing the modified footwear | All |
| *Experience with the intervention\** | Experienced discomforts while wearing the modified footwear and satisfaction with the modified footwear on a 7-point Likert scale | All |
| *Evaluation of the intervention\*<sup>#</sup>* | Experienced advantages of the modified footwear and satisfaction with the delivery process | 6, 12 |

§ For statistical analysis, participants will be dichotomized into those who experienced improvement of complaints and those who did not.
\* Only applicable for participants who filled out to have received the intervention.

#### Secondary outcome measures

Secondary outcome measures include walking pain on an 11-point NRS at 12-month follow-up. Other secondary outcomes include self-reported recovery, measured as Global Rating of Change (GROC), foot health, measured with the Foot Health Status Questionnaire (FHSQ), physical activity level, measured with the Short QUestionnaire to ASsess Health-enhancing physical activity (SQUASH) and International Physical Activity Questionnaire (IPAQ) and patient acceptable symptom state (PASS) at 6- and 12-month follow-up (Table 2).

#### Other outcomes

Other outcomes include received treatments for MTP-1 joint OA during follow-up. For people who received the intervention, the frequency and duration of wearing the modified footwear, experienced discomforts while wearing the modified footwear, satisfaction with the modified footwear, experienced advantages of the modified footwear and satisfaction with the delivery process are reported (Table 2).

### Qualitative interview study

A qualitative interview study will be performed alongside the trial with the aim to investigate the barriers and facilitators experienced by people with MTP-1 joint OA and by healthcare professionals regarding the implementation of orthopaedic modifications to off-the-shelf footwear in the treatment of MTP-1 joint OA in primary care. Identified determinants can be used in the future development of an implementation strategy that mitigates barriers and leverages facilitators, striving for successful implementation (37). Using purposeful sampling, people with MTP-1 joint OA, healthcare professionals, including orthopaedic shoe technicians and general practitioners, and representatives of health insurance companies will be interviewed until data saturation is reached. We expect to reach data saturation after 15 interviews with healthcare professionals, 15 interviews with people with MTP-1 joint OA and 5 interviews with representatives of health insurance companies. Semi-structured interviews will be conducted by a member of the research team, according to a topic guide, based on existing literature and the Consolidated Framework for Implementation Research (CFIR) (38). In the interviews, we will focus, amongst other things, on the intervention itself, including openness to, knowledge about, expectations of and experience with the intervention, as well as on the process, including patient referral, costs and the delivery process in primary care.

### Sample size

The minimal clinically important change in walking pain is 1.8 points on the 11-point NRS (39). Based on a power of 90% and a p-value of 0.05 (two-sided test) and using a conservative standard deviation (SD) of 2.8 (20), the sample size calculation resulted in 102 participants. Adjusting for crossovers and a drop-out rate of 25%, a total of 136 participants are recruited.

### Statistical analysis

An intention-to-treat approach will be used for analysis of primary and secondary outcomes. All analyses will be performed using RStudio (R Foundation for Statistical Computing, Vienna, Austria) with statistical significance set at p < 0.05. Participant characteristics and baseline values of the outcome measures will be described, using the mean (SD) for numerical data and percentages for categorical data. Additionally, per protocol sensitivity analyses will be performed for primary and secondary outcomes.

#### Effectiveness analyses

Outcome measures on each timepoint will be presented using the mean (SD) for numerical data and percentages for categorical data. For all numerical primary and secondary outcomes, treatment effects will be analysed using linear mixed models (LMM) with repeated measures. For the binomial outcomes, self-reported recovery and patient acceptability, generalised linear mixed models (GLMM) will be used with a logit link and binomial distribution. The random-effect part will consist of a subject-specific intercept to account for variability between subjects. The fixed-effect part will consist of the intervention, time and intervention×time interaction. The analysis will be adjusted for potential confounders (e.g. age, sex, symptom duration, comorbidities and BMI).

Treatment effects will be expressed in mean difference or odds ratio (OR) with corresponding 95% confidence intervals (CI) for numerical and binomial data respectively. Missing data will be handled using restricted maximum likelihood estimation within the mixed models, which provides unbiased estimates of covariance parameters under the assumption of data missing at random and does not exclude cases with partially missing data.

#### Economic evaluation

In the economic evaluation, we will relate the incremental costs to the incremental benefits of the intervention in addition to usual care, as compared to usual care alone, over a 12-month follow-up period. Both a cost-effectiveness analysis (CEA) for walking pain and a cost-utility analysis (CUA) for QALYs based on the EQ-5D-5L will be performed from a societal and healthcare perspective, according to Dutch guidelines (40). Costs will be calculated using Dutch unit costs (41).

Missing cost- and effect data will be imputed using multiple imputation according to the Multiple Imputation by Chained Equations (MICE) algorithm (42). Rubin’s rules will be used to pool the results from the different multiply imputed datasets (43).

Linear regression analyses will be used to estimate cost- and effect differences between the intervention and control group. Incremental cost-effectiveness ratios (ICERs) will be calculated by dividing the difference in mean total costs by the difference in mean total effects between the intervention and control group. Confidence intervals for the ICERs will be estimated with bias-corrected and accelerated bootstrapping with 5000 replications. The statistical uncertainty surrounding the ICERs will be graphically presented in cost-effectiveness planes and acceptability curves.

#### Qualitative data analysis

Interviews will be audio recorded and transcribed verbatim and thematically analysed, using a codebook approach (44). Themes will be generated deductively as well as inductively, using an iterative analytic process. The qualitative analysis software program MAXQDA (VERBI Software GmbH, Berlin, Germany) will be used for thematic data analysis. The first interviews will be coded independently by two researchers, after which they will compare their coded transcripts and discuss risen coding discrepancies. Disagreements will be discussed with a third researcher until consensus is reached. Once intercoder agreement is reached (i.e. no major coding discrepancies between the two researchers), remaining interviews will be coded by one researcher.

### Data monitoring and harms

Data monitoring is carried out by an independent monitor at three timepoints: after the first 15 participants are randomised, after the first participant finished follow-up and at the end-of-study. Study participation is considered a negligible risk. Serious adverse events are reported to the Medical Research and Ethics Committee (METC) via Research Portal.

### Patient and public involvement

Patient representatives from the platform Healthy with OA (45) and the Dutch Poly Osteoarthritis Society (46) have been involved in several phases of this trial. They were involved in the trial design and provided feedback on recruitment strategies and recruitment materials to verify their understandability towards the target population. In addition, patient representatives are consulted upon participant engagement and appropriate strategies to disseminate the results to people with MTP-1 joint OA. For the qualitative interview study, they will provide feedback on the topic guide for the interviews.

## ETHICS AND DISSEMINATION

The trial received ethical approval from the METC of Erasmus MC University Medical Center Rotterdam (MEC 2024-0615) in December 2024. The study protocol complies with the principles of Good Clinical Practice and the Declaration of Helsinki. The trial was prospectively registered in the Overview of Medical Research in the Netherlands (OMON) register (NL87646.078.24), which is directly linked to the International Trials Registry Platform (ICTRP).

All participants provide informed consent prior to their study participation, either written by post, or electronically using an advanced electronic signature via ValidSign (Validsign B.V., Eefde, the Netherlands). A unique numeric code is assigned to each participant, with no link to personal data. Trial data and interview data will be securely stored in a locked location for up to 15 years and will be made available upon reasonable request. Interview recordings will be deleted after transcription. Participants can additionally provide consent for their data to be stored in the OA Trial bank.

Study results will be disseminated to researchers through peer-reviewed articles in scientific journals and presentations at conferences. Study results will be disseminated to study participants, healthcare professionals and the general population through layman articles in newsletters, on websites and on social media.

## DISCUSSION

This is the first RCT to investigate the effectiveness and cost-effectiveness of a combination of multiple orthopaedic modifications to off-the-shelf footwear in addition to usual care, compared to usual care alone for people with MTP-1 joint OA. Research on footwear and orthoses in the treatment of foot OA is considered a priority by the International Foot and Ankle Osteoarthritis Consortium (22). However, research on non-surgical foot OA treatments is scarce; most articles on foot OA that were published in the last decades focused on surgical treatments or the aetiology of foot OA (5). Until now, three RCTs evaluated the effectiveness of footwear interventions in the treatment of people with MTP-1 joint OA (15) and one feasibility study compared a podiatry intervention to usual care (21). The limited evidence on the effectiveness and cost-effectiveness of footwear interventions compared to usual care and the lack of studies to a combination of multiple orthopaedic modifications to off-the-shelf footwear in the treatment of MTP-1 joint OA, underscore the relevance of our study.

Strengths and limitations of this study design need to be addressed. One strength of this study is its pragmatic design, allowing generalisation of our study findings to primary care settings across the Netherlands. In addition, performing a qualitative interview study alongside the trial will facilitate the potential implementation of orthopaedic modifications to off-the-shelf footwear in the treatment of people with MTP-1 joint OA in primary care, taking into account barriers and facilitators experienced amongst people with MTP-1 joint OA and healthcare professionals. One potential limitation of this study is that, because of the absence of diagnostic criteria for MTP-1 joint OA, we were not able to verify whether our inclusion criteria on MTP-1 joint OA diagnosis, pain and stiffness suffice in accurate selection of MTP-1 joint OA cases. Additionally, the inability of this trial to blind study participants can lead to expectation bias and may result in an overestimation of the treatment effect. However, when people with MTP-1 joint OA receive orthopaedic modifications to off-the-shelf footwear in real-world clinical practice, expectations might influence their experience and satisfaction as well, so this can be considered part of the treatment effect. Furthermore, we expect that initial expectation bias will reduce over the course of the follow-up period.

A positive study outcome would show us that orthopaedic modifications to off-the-shelf footwear are an effective treatment for people with MTP-1 joint OA. This could support healthcare decision makers to incorporate orthopaedic modifications to off-the-shelf footwear in the treatment of people with MTP-1 joint OA in primary care, which could subsequently improve patient care, reduce pain symptoms and improve quality of life. Additionally, findings from the qualitative interview study could contribute to successful implementation, taking preferences and needs of people with MTP-1 joint OA and healthcare professionals into account. Additionally, if the intervention will be shown to be cost-effective, widespread implementation of the intervention could result in reduced healthcare costs and thereby economic benefits. A negative or neutral outcome would show us that orthopaedic modifications to off-the-shelf footwear are no more effective than usual care in the treatment of people with MTP-1 joint OA. Regardless of the outcome, this trial will provide evidence on the effectiveness of an existing MTP-1 joint OA treatment and assist healthcare policy makers in the future development of clinical guidelines for MTP-1 joint OA treatment in primary care.

In conclusion, evidence on effective and cost-effective treatments for people with MTP-1 joint OA is limited and guidelines for the treatment of MTP-1 joint OA in primary care are lacking, while its estimated prevalence and burden are high. By investigating the effectiveness and cost-effectiveness of orthopaedic modifications to off-the-shelf footwear in the treatment of MTP-1 joint OA in primary care, our study will contribute to better healthcare for and improved quality of life of people with MTP-1 joint OA.

## Data Availability

No data are produced in the present study.

## ACKNOWLEDGMENTS

The authors are grateful for the contribution of patient representative Peter Froon, who was involved in the design of this trial and will stay involved during execution of the trial and dissemination of study results. Additionally, the authors would like to thank Laura van Loon, orthopaedic shoe technician, for her contribution to the intervention description and visualisation.

## AUTHORS’ CONTRIBUTIONS

MvM and SBZ conceptualised the study design and MvM acquired funding. All authors contributed to the clinical methodology and protocol refinement. SV performed project administrative tasks, under supervision of CH and MvM. SV and CH contributed to the visualisation of the intervention. SV wrote the initial draft and CH, JB, DS, MG, SK, CB, LN, SBZ and MvM critically reviewed and edited the manuscript. All authors read and approved the final manuscript.

